# Engagement with a Chatbot-based Intervention for the delivery of mailed at-home COVID-19 Testing: a Descriptive Log Analysis Study from the SCALE-UP II Trial

**DOI:** 10.64898/2026.07.08.26357530

**Authors:** Joni H. Pierce, Jiantao Bian, Tatyana V. Kuzmenko, Kimberly A. Kaphingst, Leticia Stevens, Adriana Rush, Ryzen Benson, Emerson P. Borsato, Bryan Gibson, Kensaku Kawamoto, Andy J. King, Brian Orleans, Jonathan Chipman, Tom Greene, Ray Meads, Tracey Siaperas, Shlisa Hughes, Alan Pruhs, Courtney Pariera Dinkins, Cho Y. Lam, Ryan C. Cornia, Richard L. Bradshaw, Jorie Butler, Chelsey R. Schlechter, David W. Wetter, Guilherme Del Fiol

**Affiliations:** Department of Biomedical Informatics, University of Utah, Salt Lake City, Utah, USA; Huntsman Cancer Institute, University of Utah, Salt Lake City, Utah, USA; Department of Communication, University of Utah, Salt Lake City, Utah, USA; School of Medicine Greenville, University of South Carolina, USA; University of California San Francisco, California, USA; Department of Population Health Sciences, University of Utah, Salt Lake City, Utah, USA; Association for Utah Community Health, Salt Lake City, Utah, USA; Informatics Decision-Enhancement and Analytic Sciences Center (IDEAS), VA Salt Lake City Healthcare System, Salt Lake City, Utah, USA; Division of Geriatrics, Department of Internal Medicine, University of Utah, Salt Lake City, UT, United States; Geriatrics Research, Education, and Clinical Center (GRECC), VA Salt Lake City Healthcare System, Salt Lake City, United States

## Abstract

**Background:** Promoting at-home tests (e.g., for COVID-19) using chatbots may be a novel and scalable way to improve uptake across underserved populations.

**Objective:** The objective of this study was to assess the navigational patterns (i.e., sequence of interactions) of underserved populations when using a chatbot designed to provide education on COVID-19 testing and free order access for at-home COVID-19 test kits.

**Methods:** The study was a descriptive analysis of the original data of the chatbot intervention of the SCALE-UP II trial, which compared different digital health modalities (i.e., chatbots versus simple text messages) to deliver free at-home COVID-19 test kits to minority populations in Utah.

SCALE-UP II (registration numbers NCT05533918; NCT05533359) was a multisite, pragmatic clinical trial with patients randomized in a 2×2×2 factorial design (smartphone study) to receive (1) chatbot or text messaging, (2) the option to request patient navigation, and (3) intervention frequency every 10 or 30 days. All other participants were randomized in a 2×2 factorial design (nonsmartphone study) to receive the option to request patient navigation and intervention frequency every 10 or 30 days.

Eligible patients (1) had an appointment at one of the participating community health centers (CHC) in the last 3 years, (2) were 18 years and older, and (3) had a valid cellphone number recorded in the CHC electronic health record (EHR).

The trial enrolled 2117 in the smartphone study and 31,439 in the nonsmartphone study. In the smartphone study, the proportion of participants who requested test kits in the Chatbot arm was lower than in SMS text messaging. In the nonsmartphone study, test kits was higher if they were messaged every 10 days.

Sources of funding included the National Institute on Minority Health and Health Disparities (NIMHD) of the US National Institutes of Health (NIH) grant number 5U01MD017421 and by awards from the National Cancer Institute of the NIH (P30CA042014) and the Huntsman Cancer Foundation.

**Results:** Of 1,051 patients randomized to the chatbot intervention, 309 (29%) launched the chatbot, 196 (63%) interacted with it, and 186 (60%) started the COVID-19 test kit ordering process. Among those who launched the chatbot, 170 (55%) completed a test kit order. One patient (0.3%) accessed the chatbot educational content. The median age was 51, with 66% female, 54% Latino/a, 55% uninsured, and 86% located in an urban area.

**Conclusion:** Ordering of COVID-19 test kits among underserved patients who interacted with the chatbot was high. Thus, chatbots may represent a viable approach to reach underserved populations as a part of public health response in a pandemic. All patients except one placed orders without reviewing educational content. Chatbot design should identify and minimize the number of steps for patients to achieve a specific goal.

## Introduction

During public health crises (e.g., the COVID-19 pandemic), early detection and management is central to crisis response [1]. Public health management seeks to avoid, mitigate, and control the negative impact of public health threats, especially in populations at risk for poor outcomes due to health inequities [2], [3]. Accurate and timely health education along with access to at-home COVID-19 testing may be instrumental in educating and empowering the public to assess personal risks, act in accordance with public health recommendations, and make decisions to protect themselves and those in their sphere of interaction [4].

Digital health interventions such as text messaging and chatbots are promising approaches for providing health education and creating a mechanism to provide access to at-home testing [5], [6]. Chatbots are computer programs designed to simulate human conversation [7], [8]. Systematic reviews have shown that interventions using chatbots are effective for a wide range of clinical problems, including treatment and monitoring, health services support, diagnostics, education, and lifestyle and behavioral change [9–12] [13] [14]. Chatbots can reach users where health care is limited and offer a range of benefits such as convenience, scalability, personalization, and ease of use controlled by the patient [15]. According to a scoping review, several chatbots were developed during the COVID-19 pandemic to provide information, personal risk assessment and triage, exposure notification and monitoring, and infection tracking [16], [17,18]. For example, Perez-Ramos et al. found that a COVID-19 chatbot to be a feasible strategy to provide COVID-19 vaccine information and access to historically marginalized groups [19].

We found a mix of scoping reviews and individual studies focused on chatbots for underserved populations and most studies were dated 2020 or later. Some studies were mainly designed for women and adolescent girls and focused on mental health, sexual health, violence prevention, and COVID-19 information and vaccination promotion [20]. Other studies focused on improving health equity and access to care, mental health, and vaccine promotion [21], [22,23], [24] [25], [26], [27],[28,29].

A few studies reported user concerns related to the impersonal nature of a chatbot [30], [31]. A scoping review by Xue mapped studies focused on general healthcare chatbots and found mental health, health promotion and behavior change as key theme in the literature. They noted suicidal thought intervention in 44% of the included studies [32]. Similarly, Wilson noted 30% of the studies included in their scoping review focused on mental health and 20% on COVID-19 related topics [33].

The objective of this study was to assess patients’ navigational patterns (i.e., sequence of interactions) when using a chatbot designed to provide education on COVID-19 testing and to order free at-home COVID-19 test kits in underserved populations. The study was a descriptive analysis of the original data from the SCALE-UP II, a pragmatic randomized controlled trial that investigated different digital health interventions promoting access to at-home COVID-19 testing among patients who receive care at low resource community health centers (CHCs) across the state of Utah [6].

## Methods

### Study Population

The study population was a subset of the SCALE-UP II trial population, which targeted underserved populations in Utah. We included individuals who were randomized to the chatbot arm and opened the chatbot at least once during the course of the trial, from December 19, 2022 to August 28, 2023. The inclusion criteria for SCALE-UP II was 18 years of age or older, had English or Spanish as the preferred language recorded in the electronic health record (EHR), had a valid cellphone number recorded in the EHR, and received care at one of 12 clinics within 3 safety net CHCs in Utah. Participating CHCs serve over 39,000 patients per year; 46% of patients are Hispanic/Latino, 54% uninsured, and three of the 12 clinics are located in rural areas. Rural or urban status was determined based on guidelines defined by the Federal Office of Rural Health Policy (FORHP) [34].

### Chatbot content design

The chatbot script was created in both English and Spanish, and the language of the chatbot was automatically selected based on the patient’s preference recorded in the clinic’s EHR. An interdisciplinary team of researchers with expertise in digital health, health communication, sociotechnical methods and biomedical informatics met on a weekly basis to discuss the design of the chatbot script. Chatbot script design was guided by health communication theories such as the extended parallel process model (EPPM) [35]; recommendations from sources including the US Centers for Disease Control, Utah Department of Health and Human Services, and COVID-19 test kit product manufacturers; and formative interviews and surveys with members of the target population. The resulting script started with an introduction explaining how to use the chatbot (Figure 1a), followed by a brief explanation about testing along with the option to order a test (Figure 1b) or to review educational content (Figure 1d). Educational content included topics such as (1) why, when, and how to use a test; (2) COVID-19 symptoms; and (3) what to do if a test is positive (e.g., quarantine, when and where to find treatment). Patients who placed an order were asked to confirm their mailing address available in the clinic’s EHR (Figure 1c). If the address was incorrect, the patient had the opportunity to correct it (Figure 2). The chatbot software tracked detailed user interactions with timestamps through an audit log, which is the source of the data that was used for this study.

**Figure 1.**
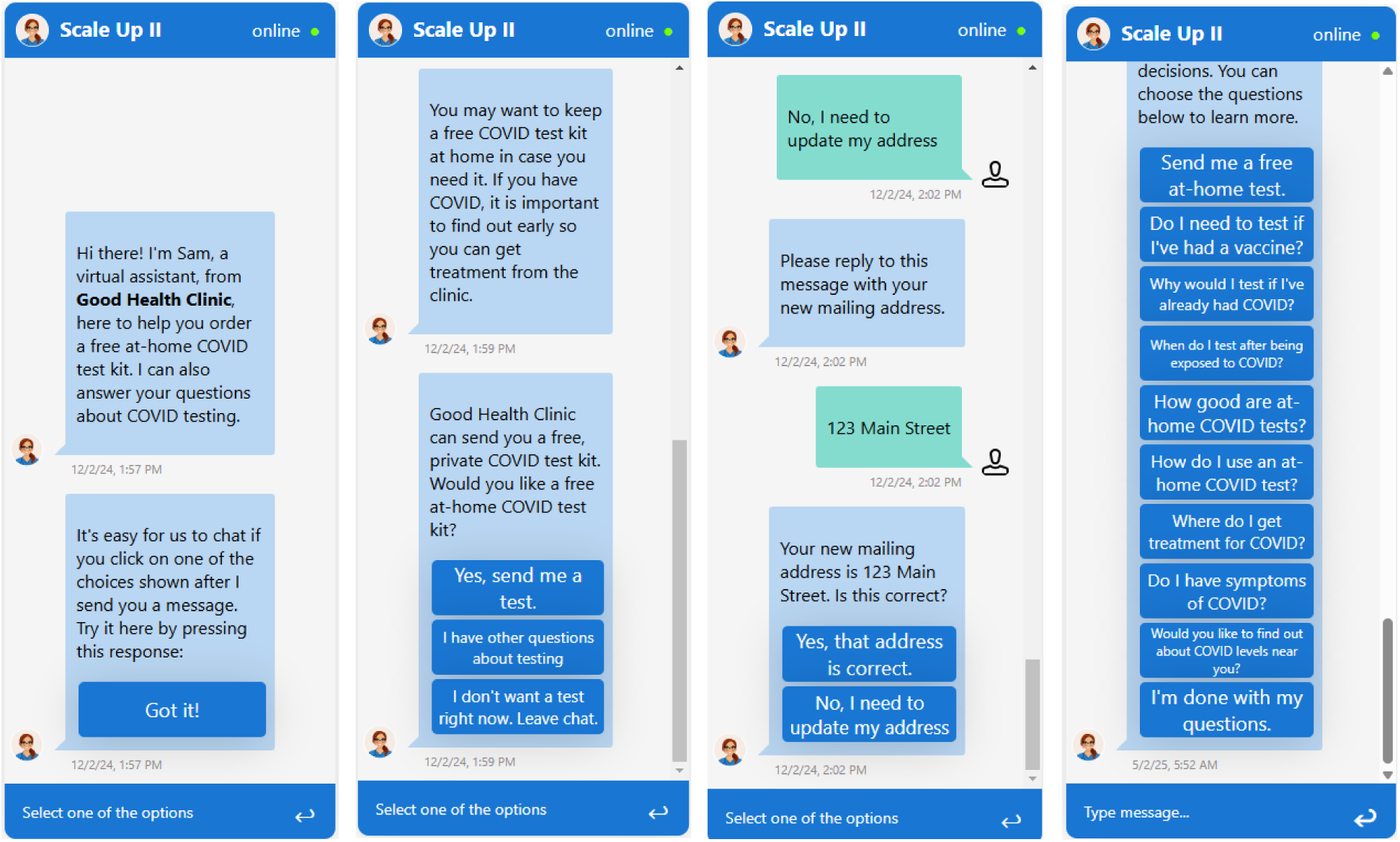
COVID-19 at-home test kit chatbot interaction screenshots. (1a) Introduction; (1b) Test offer and option to review education content; (1c) Address verification; (1d) Educational topics. (Dec 2022 – Aug 2023). A secondary data analyses of SCALE-UP II, a pragmatic randomized controlled trial that investigated different digital health interventions promoting access to at-home COVID-19 testing among patients who receive care at low resource community health centers (CHCs) across the state of Utah.

**Figure 2.**
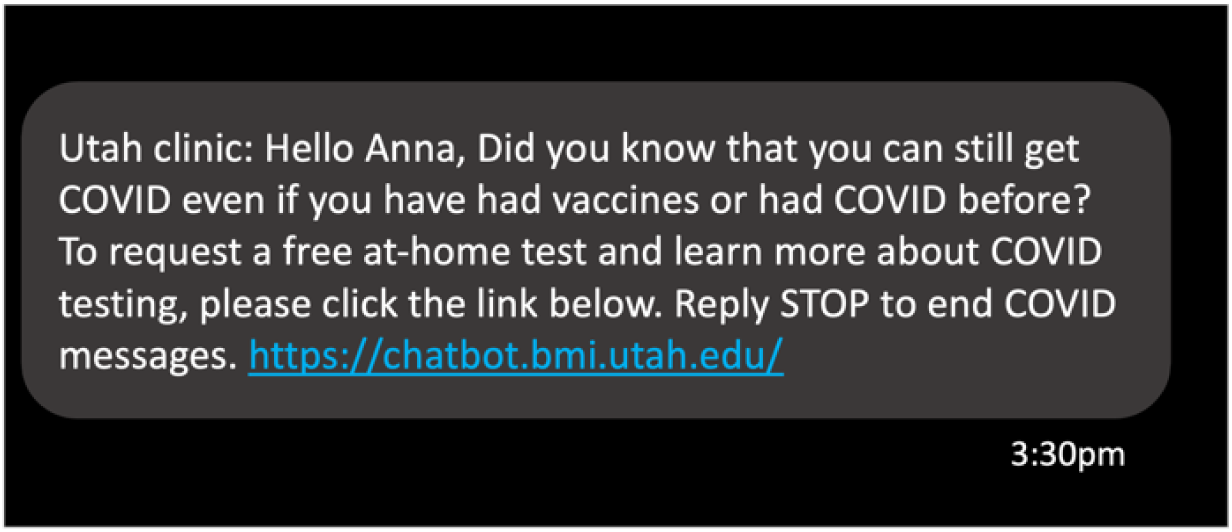
Text message screenshot with a hyperlink to launch the COVID-19 education and at-home test kit ordering chatbot. (Dec 2022 – Aug 2023). A secondary data analyses of SCALE-UP II, a pragmatic randomized controlled trial that investigated different digital health interventions promoting access to at-home COVID-19 testing among patients who receive care at low resource community health centers (CHCs) across the state of Utah.

All adult patients who had a primary care appointment in the last three years at one of the study sites were automatically enrolled in the SCALE-UP II trial. All patients received a text messaging asking whether they had a smartphone with access to the internet. Those who replied “Yes” were randomized to a text messaging versus a chatbot arm. Patients randomized to the chatbot arm were able launch the chatbot by clicking a hyperlink sent via a text message on behalf of the patient’s clinic (Figure 2). Text messages were sent every 10 or 30 days for a total of 6 or 3 messages respectively.

Chatbot users were not able to provide open-ended, free-text questions. The chatbot used a rule-based designed in which all content was scripted, with pre-defined navigation options and content.

### Data cleaning and processing

Chatbot log data were exported as comma-separated files (CSV) files and then cleaned for analysis. The study has no missing data. All cleaning and analyses were performed within a Python Jupyter notebook using Python (version 3.12.7), with Pandas (version 2.2.2) and NumPy (version 2.0.0). Chatbot log data were linked with demographic data from CHC EHRs. Next, chatbot navigation events were classified into two overarching navigational categories: test ordering steps (i.e., starting a test order, address confirmation, address correction) and use of educational content.

### Data analysis and interpretation

Descriptive statistics included time spent interacting with the chatbot, and counts and frequencies of chatbot navigation events.

### Ethical considerations

SCALE-UP II was approved by the University of Utah Institutional Review Board (protocol 00150669) and follows the CONSORT (Consolidated Standards of Reporting Trials) reporting guidelines [36]. The trial was registered with Clinicaltrials.gov (NCT05533359 for the Smartphone study and NCT05533918 for the Nonsmartphone study). We prospectively registered the study on 9/7/22 with the first participant enrolled on 12/19/22. https://clinicaltrials.gov/study/NCT05533918?tab=history&a=3#version-content-panel. As a pragmatic trial with minimal risk, the University of Utah Institutional Review Board approved a waiver of consent for enrollment, thus consent was not required. Therefore, all eligible participants were automatically enrolled in and blinded to the study. All participant identification data were anonymized and aggregated; therefore, no identification of individual participants is possible based on the manuscript, supplemental materials or other associated data. No compensation was offered to the SCALE-UP II participants.

## Results

Among the 1,051 patients randomized to the chatbot intervention in SCALE-UP II, 309 (29%) opened the chatbot at least once and were included in the chatbot log analysis study. Table 1 describe the demographics of study participants (N=309). The median age of patients was 51 years (range 18 to 90), 204 (66%) were female, 108 (35%) identified as Non-Latino White, 166 (54%) identified as Latino, 147 (48%) indicated Spanish as their preferred language in the EHR, 267 (86%) lived in an urban area and 170 (55%) were uninsured.

**Table 1.** COVID-19 chatbot at-home test kit ordering. Study population demographics. (Dec 2022 – Aug 2023). A secondary data analyses of SCALE-UP II, a pragmatic randomized controlled trial that investigated different digital health interventions promoting access to at-home COVID-19 testing among patients who receive care at low resource community health centers (CHCs) across the state of Utah.

|  |  |
| --- | --- |
| N | 309 |
| **Age (in years)** |  |
| *Mean (SD%)* | 51 (15%) |
| **Sex, No. (%)** |  |
| *Female* | 204 (66%) |
| *Male* | 104 (34%) |
| *Unknown* | 1 (.3%) |
| **Race/Ethnicity, No. (%)** |  |
| *American Indian/Alaska Native* | 1 (.3%) |
| *Asian* | 1 (.3%) |
| *Black/African American* | 2 (.6%) |
| *Hawaiian/Pacific Islander* | 1 (.3%) |
| *Hispanic or Latino/a* | 166 (54%) |
| *Multi-race* | 3 (.9%) |
| *White* | 244 (79%) |
| *Unknown* | 28 (11%) |
| **Urban vs. Rural, No. (%)** |  |
| *Urban* | 267 (86%) |
| **Language, No. (%)** |  |
| *English* | 149 (48%) |
| *Spanish* | 147 (48%) |
| *Other* | 4 (1%) |
| *Unknown* | 9 (3%) |
| **Insurance type. (%)** |  |
| *Private* | 78 (25%) |
| *Public* | 61 (20%) |

Of the 309 patients who launched the chatbot, 186 (60%) started a test kit order, and 170 (55%) completed the test order. Of 34 (11%) patients who reported their address was not correct, 24 (8%) updated their address. One patient explored the educational content (Figure 3).

**Figure 3.**
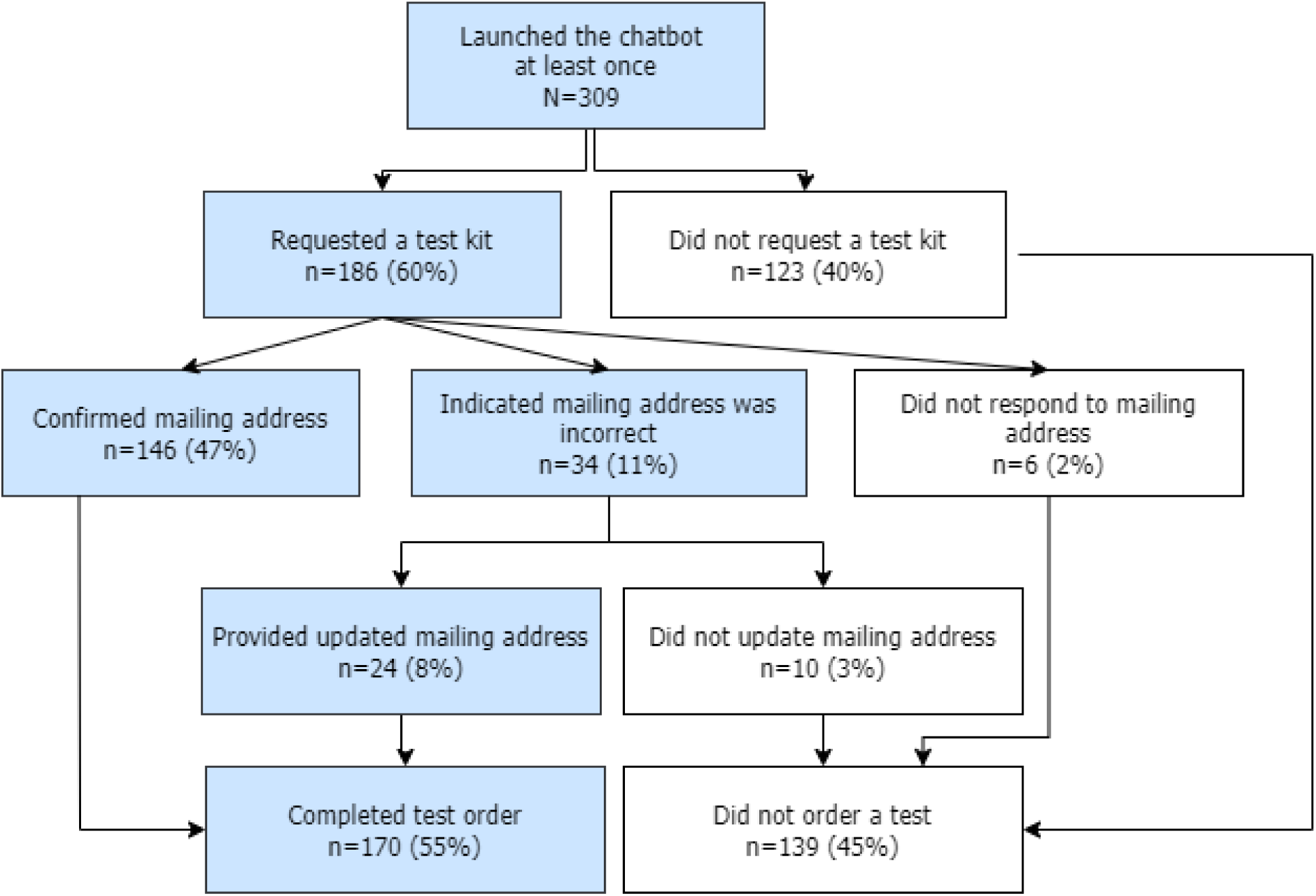
**COVID-19 chatbot at-home test kit ordering process. (**Dec 2022 – Aug 2023). A secondary data analyses of SCALE-UP II, a pragmatic randomized controlled trial that investigated different digital health interventions promoting access to at-home COVID-19 testing among patients who receive care at low resource community health centers (CHCs) across the state of Utah.

### Navigation with the chatbot

Among 309 patients who launched the chatbot, 113 (37%) never interacted with the chatbot beyond the brief introduction explaining how to use the chatbot. Among the 196 patients who interacted with the chatbot, 186 (95%) started a test order and 170 (87%) completed the ordering process.

Among patients who interacted with the chatbot, the median session time was 70 seconds for those who started a test order versus 113 seconds among those who did not order (mean = 135 sec vs. 337 sec). Among those who started a test kit order, the median session time was 83 seconds for those who did not complete the order (i.e., did not confirm or correct their address) and 66 seconds for those who completed the order (mean = 136 sec vs. 135 sec).

### COVID-19 chatbot at home test ordering process

## Discussion

We assessed the navigational patterns of using a chatbot to deliver mailed at-home COVID-19 test kits among patients who receive care at safety net CHCs. Overall, patients used the chatbot to request COVID-19 test kits without accessing any of the educational components. More than half of those who launched the chatbot completed an order for an at-home COVID-19 test kit. This implies that chatbots may be a viable outreach method for populations in low resource settings. All but one patient who ordered test kits did so without engaging with optional educational chatbot content. Consequently, Chatbot design should minimize the number of steps for patients to achieve a goal. Previous studies on COVID-19 chatbots have focused largely on issues such as symptom checking and vaccination eligibility [16,17], [18], [12]. This study adds to the existing body of research by analyzing engagement patterns of patients from low resource settings with a scalable chatbot-based intervention aimed at delivering mailed at-home COVID-19 tests.

Over half of patients (55%) who launched the chatbot completed the ordering process for a test kit, which is relatively high when considering other published studies evaluating chatbots for diagnostic testing. In contrast to other published studies, this is a high engagement response [37], [38]. For example, in a trial that investigated chatbots for genetic testing of familial cancer, 31% of patients launched the chatbot and 26% completed pre-test genetic test education [37]. Another randomized controlled trial compared a digital health intervention using WhatsApp messenger versus usual care for colorectal cancer screening using fecal immunochemical tests (FIT). Test pick up and completion rates increased significantly through the introduction of the WhatsApp messenger intervention [38]. The implication based on the findings of this study and others, is that chatbots may represent a reasonable modality to reach underserved populations for the promotion of public health programs.

In our study only one patient accessed educational content, which may indicate that information needs were low and felt to be not necessary prior to ordering a test. This is consistent with behavioral economics as patients exhibited expediency in obtaining a free at-home COVID-19 test [39]. This implies that chatbot designers should thoughtfully research and design content components to ensure a chatbot contains the minimum effective content components.

Launch timing is also a key consideration for an interventional chatbot. Given our study timing in relation to the course of the pandemic, prior exposure to other sources of information may have led to reducing and shifting patients’ perceived information needs about COVID-19 testing [40]. By December 2022 when the SCALE-UP II trial launched, education campaigns had saturated both broadcast and social media channels, school and community venues, and clinical settings [41], [42]. A high level of information saturation may have influenced the interest level in the COVID-19 educational content as the public began to exhibit COVID-19 messaging fatigue and information overload [42], [43]. According to Sun et al., COVID-19 social media messaging fatigue was found to increase COVID-19 message avoidance [44].

### Comparison with Prior Work

We found a mix of scoping reviews and individual studies focused on chatbots for underserved populations. Topics ranged from mental health, sexual health, violence prevention, and COVID-19 information and vaccination promotion, improving health equity and access to care [20], [21], [22,23], [24] [25], [26], [27],[28,29].

We were unable to find any published studies to compare our findings related to chatbot navigational patterns within underserved populations. Accordingly, our study expands the existing literature by elucidating the navigational patterns of a chatbot designed to support at-home COVID-19 testing in underserved populations. Aspects of our findings may translate to other public health scenarios involving the promotion of home based testing.

Our study suggests digital health interventions in the form of chatbots could play an important role in public health emergencies/situations to reach underserved populations to promote access to care.

### Implications for Chatbot Design

A sizable portion of patients (37%) who launched the chatbot never interacted beyond the brief introduction explaining how to use the chatbot. On the other hand, the vast majority (87%) of those who interacted past the brief introduction completed a test kit order. It is possible that the brief introduction became a barrier to interaction and caused attrition. Therefore, chatbot design should consider minimizing the number of steps needed for patients to achieve a specific goal.

The timing of outreach via chatbots for the delivery of educational content should be explored in future studies. Furthermore, information needs and communication goals require independent consideration when determining the scope of a chatbot design. If deemed appropriate, a pre-test chatbot could offer brief educational content such as a short video explaining the benefits of testing, followed by the ordering workflow. Educational content could also be offered via chatbot after a patient receives a test kit to explain how to use a test and what to do if a test result is positive.

### Limitations

This study offers important insights into navigational patterns of a chatbot intervention but also has limitations. First, the study findings reflect chatbot use patterns at a specific point in the context of a pandemic. Consequently, it is possible that the study findings will not generalize to other public health contexts such as at the inception of a pandemic or routine preventive care testing, such as cancer screening. Furthermore, the COVID-19 pandemic became highly politicized and plagued by mis/dis/mal information[45], [46], [47], [48]. More research targeting other at-home testing scenarios (e.g., cancer screening, other infectious diseases) should be undertaken. Second, log data reflects user behavior, however it does not inform as to the motivation for the behavior. Due to the pragmatic design, the SCALE-UP II trial did not collect individual patient characteristics beyond demographics that may drive these behaviors. However, we were able to look at use of the chatbot in a population context. Last, the study population reflects historically marginalized populations in Utah and may not generalize to populations in other geographic locations.

### Conclusion

This study found that the majority of patients who interacted with the chatbot completed an order for an at-home COVID-19 test. Users did not access the educational content which suggests users navigational patterns were aimed at ordering but not information seeking. A chatbot-based intervention resulted in a relatively high rate of ordering among patients receiving care at safety net community health centers. This underscores the viability of new modalities such as chatbots as a digital health intervention as a modality to reach underserved populations for the promotion of at-home testing. This supports the democratization of care and health equity by promoting early awareness and access to testing resources in the context of public health events.

## Use of AI

AI technologies were not used in for any stage of the research process or the preparation of the manuscript.

## Data Availability

The datasets generated and analyzed during this study are available from the corresponding author on reasonable request.

## Authors’ Contributions

All authors provided contributions to the conceptualization of the study. JB, TVK, BO and JC conducted data curation and verification. JB and TVK conducted formal analysis. JP, GDF and TVK performed data interpretation. EPB, RCC, and RLB developed the software infrastructure and digital health interventions. All authors contributed to the development of the manuscript.

## Abbreviations

CHC: community health center
CONSORT: Consolidated Standards of Reporting Trials
CVS: comma-separated values
FIT: fecal immunochemical test
EHR: electronic health record
EPPM: extended parallel processing model
NIH: national institutes of health
HIMHD: national institute on minority health and health disparities
RCT: randomized clinical trial

## References

1. Rai P, Kumar BK, Deekshit VK, Karunasagar I, Karunasagar I. Detection technologies and recent developments in the diagnosis of COVID-19 infection. Appl Microbiol Biotechnol. 2021;105: 441–455.

2. Khanijahani A, Iezadi S, Gholipour K, Azami-Aghdash S, Naghibi D. A systematic review of racial/ethnic and socioeconomic disparities in COVID-19. Int J Equity Health. 2021;20: 248.

3. Schaffer DeRoo S, Torres RG, Ben-Maimon S, Jiggetts J, Fu LY. Attitudes about COVID-19 testing among Black adults in the United States. Ethn Dis. 2021;31: 519–526.

4. Peeling RW, Heymann DL, Teo Y-Y, Garcia PJ. Diagnostics for COVID-19: moving from pandemic response to control. Lancet. 2022;399: 757–768.

5. Del Fiol G, Orleans B, Kuzmenko TV, Chipman J, Greene T, Martinez A, et al. SCALE-UP II: protocol for a pragmatic randomised trial examining population health management interventions to increase the uptake of at-home COVID-19 testing in community …. BMJ open. 2024;14: e081455.

6. Del Fiol G, Kuzmenko TV, Orleans B, Chipman J, Greene T, Meads R, et al. SCALE-UP II: Pragmatic Randomized Trial Examining Population Health Management Interventions to Increase the Uptake of At-Home COVID-19 Testing Among Community Health Centers …. Available: https://scholar.google.com/citations?view_op=view_citation&hl=en&citation_for_view=helFq1gAAAAJ:R3hNpaxXUhUC

7. Adamopoulou E, Moussiades L. An Overview of Chatbot Technology. Iliadis L, Pimenidis E, editors. Cham, Switzerland: Springer International Publishing; 2020.

8. Martinengo L, Jabir AI, Goh WWT, Lo NYW, Ho M-HR, Kowatsch T, et al. Conversational agents in health care: Scoping review of their behavior change techniques and underpinning theory. J Med Internet Res. 2022;24: e39243.

9. Luk TT, Lui JHT, Wang MP. Efficacy, Usability, and Acceptability of a Chatbot for Promoting COVID-19 Vaccination in Unvaccinated or Booster-Hesitant Young Adults: Pre-Post Pilot Study. J Med Internet Res. 2022;24: e39063.

10. Okonkwo CW, Amusa LB, Twinomurinzi H. COVID-Bot, an Intelligent System for COVID-19 Vaccination Screening: Design and Development. JMIR Form Res. 2022;6: e39157.

11. Rizzato Lede DA, Inda D, Rosa JM, Zin Y, Tentoni N, Médici MM, et al. Tana, a Healthcare Chatbot to Help Patients During the COVID-19 Pandemic at a University Hospital in Argentina. Stud Health Technol Inform. 2022;290: 301–303.

12. Siedlikowski S, Noël LP, Moynihan SA, Robin M. Chloe for COVID-19: Evolution of an Intelligent Conversational Agent to Address Infodemic Management Needs During the COVID-19 Pandemic. J Med Internet Res. 2021;23: e27283.

13. Rathnayaka P, Mills N, Burnett D, De Silva D, Alahakoon D, Gray R. A Mental Health Chatbot with Cognitive Skills for Personalised Behavioural Activation and Remote Health Monitoring. Sensors (Basel). 2022;22. doi:10.3390/s22103653

14. Tudor Car L, Dhinagaran DA, Kyaw BM, Kowatsch T, Joty S, Theng Y-L, et al. Conversational agents in health care: Scoping review and conceptual analysis. J Med Internet Res. 2020;22: e17158.

15. Pereira J, Díaz Ó. Using health chatbots for behavior change: A mapping study. J Med Syst. 2019;43: 135.

16. Almalki M, Azeez F. Health chatbots for fighting COVID-19: A scoping review. Acta Inform Med. 2020;28: 241–247.

17. Amiri P, Karahanna E. Chatbot use cases in the Covid-19 public health response. J Am Med Inform Assoc. 2022 [cited 22 Mar 2022]. doi:10.1093/jamia/ocac014

18. Miner AS, Laranjo L, Kocaballi AB. Chatbots in the fight against the COVID-19 pandemic. NPJ Digit Med. 2020;3: 65.

19. Perez-Ramos JG, Leon-Thomas M, Smith SL, Silverman L, Perez-Torres C, Hall WC, et al. COVID-19 vaccine equity and access: Case study for health care chatbots. JMIR Form Res. 2023;7: e39045.

20. Hazhir S, Langarizadeh M, Bahariniya S, Valizadeh Laktarashi H, Hami A, Fatemi Aghda SA. The development and use of chatbots in enhancing health care access for underserved and vulnerable populations: a scoping review. BMC Public Health. 2025;26: 410.

21. Hu Y, Krueger A. Using digital technology to advance health equity. Health Equity: Strategies for Action. 2025. Available: https://books.google.com/books?hl=en&lr=&id=vAifEQAAQBAJ&oi=fnd&pg=PA135&dq=health+equity+and+chatbots&ots=mQxwBjhGwm&sig=ewtmbLE8-L052d8I_RXxDvjdL7E

22. Tzelios C, Contreras C, Istenes B, Astupillo A, Lecca L, Ramos K, et al. Using digital chatbots to close gaps in healthcare access during the COVID-19 pandemic. Public Health Action. 2022;12: 180–185.

23. Sezgin E, Kocaballi AB, Dolce M, Skeens M, Militello L, Huang Y, et al. Chatbot for social need screening and resource sharing with vulnerable families: Iterative design and evaluation study. JMIR Hum Factors. 2024;11: e57114.

24. Morias A, Renmans D. Conversational agents and equity in health care systems: a scoping review. J Health Equity. 2026;3. doi:10.1080/29944694.2025.2606723

25. Weeks R, Cooper L, Sangha P, Sedoc J, White S, Toledo A, et al. Chatbot-Delivered COVID-19 Vaccine Communication Message Preferences of Young Adults and Public Health Workers in Urban American Communities: Qualitative Study. J Med Internet Res. 2022;24: e38418.

26. Frietze GA, Mancera BM, Kenney MJ. COVID-19 testing, vaccine perceptions, and trust among Hispanics residing in an underserved community. Int J Environ Res Public Health. 2023;20: 5076.

27. Le Bonniec A, Sauvaget C, Lucas E, Nassiri A, Selmouni F. Design and validation of a chatbot-based cervical cancer screening decision aid for women experiencing socioeconomic disadvantage: User-centered approach study. JMIR Cancer. 2025;11: e70251.

28. Revathi, Priyanka S, Priyadharshini, Nn N, Visalachi, Sumathi P. AI-driven approaches to enhancing mental wellbeing and stress relief. 2025 International Conference on Multi-Agent Systems for Collaborative Intelligence (ICMSCI). IEEE; 2025. pp. 925–931.

29. Elyoseph Z, Gur T, Haber Y, Simon T, Angert T, Navon Y, et al. An ethical perspective on the democratization of Mental Health with generative AI. JMIR Ment Health. 2024;11: e58011.

30. Moore AA, Ellis JR, Dellavalle N, Akerson M, Andazola M, Campbell EG, et al. Patient-facing chatbots: Enhancing healthcare accessibility while navigating digital literacy challenges and isolation risks-a mixed-methods study. Digit Health. 2025;11: 20552076251337321.

31. Brennan K. Chatbots in sexual and reproductive health: bridging the divide in accessibility and equity. Contraception. 2026;157: 111199.

32. Xue J, Zhang B, Zhao Y, Zhang Q, Zheng C, Jiang J, et al. Evaluation of the current state of chatbots for digital health: Scoping review. J Med Internet Res. 2023;25: e47217.

33. Wilson L, Marasoiu M. The development and use of chatbots in public health: Scoping review. JMIR Hum Factors. 2022;9: e35882.

34. HRSA. Federal Office of Rural Health Policy (FORHP). 2025. Available: https://www.hrsa.gov/about/organization/bureaus/forhp (accessed 5/4/2025)

35. Witte K. Putting the fear back into fear appeals: The extended parallel process model. Commun Monogr. 1992;59: 329–349.

36. Hopewell S, Chan A-W, Collins GS, Hróbjartsson A, Moher D, Schulz KF, et al. CONSORT 2025 statement: updated guideline for reporting randomized trials. Nat Med. 2025;31: 1776–1783.

37. Chavez-Yenter D, Kimball KE, Kohlmann W, Lorenz Chambers R, Bradshaw RL, Espinel WF, et al. Patient interactions with an automated conversational agent delivering pretest genetics education: Descriptive study. J Med Internet Res. 2021;23: e29447.

38. Lam TYT, Wu PI, Tang RSY, Luk AKC, Ng S, Sung JJY. Mobile messenger-initiated reminders improve longitudinal adherence in a community-based, opportunistic colorectal cancer screening program: A single-blind, crossover randomized controlled study. Cancer. 2021;127: 914–921.

39. Reed KL, Harvey EM, Everly CJ. The intersection of Behavioral Economics and the general medicine literature. Am J Med. 2021;134: 1350–1356.e2.

40. Mangono T, Smittenaar P, Caplan Y, Huang VS, Sutermaster S, Kemp H, et al. Information-seeking patterns during the COVID-19 pandemic across the United States: Longitudinal analysis of Google Trends data. J Med Internet Res. 2021;23: e22933.

41. Breyton M, Schultz É, Smith A “ben,” Rouquette A, Mancini J. Information overload in the context of COVID-19 pandemic: A repeated cross-sectional study. Patient Educ Couns. 2023;110: 107672.

42. Mohammed M, Sha’aban A, Jatau AI, Yunusa I, Isa AM, Wada AS, et al. Assessment of COVID-19 information overload among the general public. J Racial Ethn Health Disparities. 2022;9: 184–192.

43. Jia X, Ahn S, Carcioppolo N. Measuring information overload and message fatigue toward COVID-19 prevention messages in USA and China. Health Promot Int. 2023;38. doi:10.1093/heapro/daac003

44. Sun J, Lee SK. “No more COVID-19 messages via social media, please”: the mediating role of COVID-19 message fatigue between information overload, message avoidance, and behavioral intention. Curr Psychol. 2023; 1–15.

45. Bolsen T, Palm R. Politicization and COVID-19 vaccine resistance in the US. Progress in molecular biology and translational science. 2022;188: 81–100.

46. Hart PS, Chinn S, Soroka S. Politicization and polarization in COVID-19 news coverage. Sci Commun. 2020;42: 679–697.

47. Hardy LJ, Mana A, Mundell L, Neuman M, Benheim S, Otenyo E. Who is to blame for COVID-19? Examining politicized fear and health behavior through a mixed methods study in the United States. PLoS One. 2021;16: e0256136.

48. Newport F. The partisan gap in views of the coronavirus. Gallup, May. 2020;15.

